# Neonatal death: supporting bereaved mothers

**DOI:** 10.1101/2020.07.11.20151431

**Authors:** Alia Embaireeg, Amal KA Ayed, Mariam Khader Ayed

**Author notes:** **Corresponding Author:** Mariam Khader Ayed, Department of Neonatology, Farwaniya Hospital, Subah An Nasser, Kuwait City, Postal code-81400, Kuwait. Coauthor (Alia Embaireeg), (Amal KA Ayed).

## Abstract

**Background:** Death of a child is a devastating experience for parents, owing to which parents may show dissatisfaction towards medical care or suffer from intense and prolonged grief. The objective of the present study was to explore the needs of bereaved mothers after the death of their infant.

**Methods:** The present study was a descriptive qualitative design that consisted of 10 mothers who have been bereaved in the past year. All mothers were recruited from the registry. Data were obtained through an unstructured single interview and analyzed using conventional content analysis.

**Results:** The current study demonstrated that parents had the same needs despite their different backgrounds, socioeconomic standards, and religious beliefs. Their needs were based on several main points; 1) Lack of sensitivity/method in delivering the news, 2) Bonding with the child; 3) Safety and family support; 4) Providing understanding and meaning; 5) Ability to express emotions.

**Conclusions:** The key components that should be considered to support the bereaved parents include honesty, information, choices, and timing. The present study provided a comprehensive view of the overall experiences of the parents with neonatal death and the ability to give guidance to the healthcare providers.

## Background

The death of a child is a devastating experience and profound loss for a mother that is difficult to acknowledge. At this time, a mother’s need is to appropriately grieve for the loss of her child. The death of a baby is especially difficult to endure because mothers envision an entire lifetime for their children from the moment of confirmation of pregnancy. Moreover, their expectations and vision that were built over time are shattered with the death of their baby; from that moment an entire future is lost with the mother’s sense of parenthood [1-3]. During the perinatal period mother’s will have a lingering sense that further loss might be coming, This type of grief in which mother’s start to experience; when she is informed that her unborn child may not survive in utero or during or after delivery. The baby that survives delivery but is critically ill brings a combination of emotional reactions to the birth of the infant, Joy in welcoming a new life and fear pending loss which has psychological, cognitive, physiological and behavioural manifestations [1, 2]. In Valizadeh’s study parents reported grief reactions such as feelings of sadness, anger, difficulty in sleeping and appetite, preoccupation thinking, irritability and guilt [3]. This stigmatization of perinatal loss intensifies the grieving process of mother’s that lead to self harm, feelings of disbelief, detachment, despair and disorganization [4]. Add to that the memories that the mother come across in the NICU (Mix of new unfamiliar faces, Machines, Tubes, high noise level, Lack of privacy, loss of parent caretaking role and periods of separation) have shown to have a negative effect for years after the baby’s death [5]. In other words unexpected events alter the role of motherhood.

Maternal grief may be compounded by social stigma, blame, and marginalization. Practices of isolating women and their newborns and a perception that the newborn is not a person contribute to suboptimal care for parents when a baby dies [4]. Kowalski states that perinatal death results in multiple losses to parents, including the loss of a significant person, the loss of some aspect of the self, the loss of external object, the loss of a stage of life, the loss of a dream, and the loss of creation [6]. Culturally, a couple whose first pregnancy ends in a loss could not complete the rite of passage into parenthood, which symbolizes adult status [3]. Therefore, it has been documented that, compared with other types of bereavement, parental grieving is particularly intense, complicated, and long lasting, with major and unparalleled symptom fluctuations over time. Grieving mother’s experience intense emotions of physical pain, Self loathing, guilt, psychiatric symptoms are also common depression, anxiety, PTSD, traumatic grief which has led to further isolation [7].

An important part of parental bereavement and how they face the imminent future ahead of them is the relationship they have built over time with their healthcare professional from the moment they have conceived. Physicians do not wish to take hope away from the parents, especially when it comes to informing the parents about the death of their child/infant; it is an unpleasant task. At the same time, they may fear the unpredictability of both the reaction and the intense emotional response of the mother. Bad news often must be delivered in settings that are not conducive to such intimate conversations. The hectic pace of clinical practice may force a physician to deliver bad news grieve with little forewarning or when other responsibilities are competing for the physician’s attention [5, 6]. HCP may be able to anticipate potential sources of stress and intervene by having the parents involved in the care of their baby to decrease the long term grief [8, 9].

Little is known in our region and particularly Kuwait about the psychological needs of bereaved mothers, possibly due to the inadequate training and reluctance to discuss death issues. Our goal is to evaluate the needs of bereaved mothers in Kuwait to be able to acknowledge this issue more effectively.

## Methods

This study was conducted at tertiary neonatal intensive care unit (NICU), Kuwait. The annual total deliveries are approximately 7500 per year with total admissions to the NICU being approximately 450 per year. Neonatal mortality rate is about 1.9/1000 live birth. The main causes of death are multiple congenital anomalies, extreme prematurity and its related complications, birth asphyxia and sepsis. In this study, convenient samples of 12 mothers who have experienced a neonatal death in the last 12 months were identified from a mortality registry log. Ten mothers consented to the interview. They were asked four open-ended questions in addition to demographic questions. All interviews were conducted by the first and second author. Eight mothers participated in audio-recorded interviews lasting 45 to 60 minutes. Two mothers preferred telephonic interview. The participants represented 13 children who died from extreme prematurity (n=5), chromosomal anomalies (n=2), sepsis with multi-organ failure (n=2), and congenital diaphragmatic hernia (n=1).

A conventional approach content analysis method was used to analyse the response and identify themes within mothers’ responses. Data were analysed based on the method proposed by Lundman and Graneheim [10, 11]. Interviews were started by asking the mothers to share their NICU journey, starting from delivery to NICU admission. Later, the interview focused on the NICU experiences from the time of diagnosis until the point where they learned that their child was critically ill. The unit of analysis within each response was the meaning of the phrases. Each meaning was then combined to a common theme. Two authors (MA and AA) separately analysed and categorized participants’ responses. For inter rater agreement between the authors on each theme we calculated the Cohen Kappa coefficient.

The study was approved by IRB from the Ministry of Health of Kuwait. The informed consent was obtained verbally from all individual participants included in the study.

### Questions

1. Can you share with us your NICU journey, from delivery to NICU admission?
2. How did you find out that your child was dying?
3. Following the death of your child, how did you cope?
4. Would you suggest for better ways for us to deliver the diagnosis, death of the child, and how to support the parents after the death of their child?

## Results

Mothers’ demographic data are shown in Table 1. From the interviews that were sought, five emerging themes were generated to describe mother’s bereavement after neonatal death (Table 2). The Cohen Kappa coefficient was 0.91 for theme 1 (bonding with their child), 0.95 for theme 2 (explaining), 0.89 for theme 3 (sensitivity while delivering the news), 0.93 for theme 4 (ability to express emotions) and 0.88 for theme 5 (safety and family support).

**Table 1:**
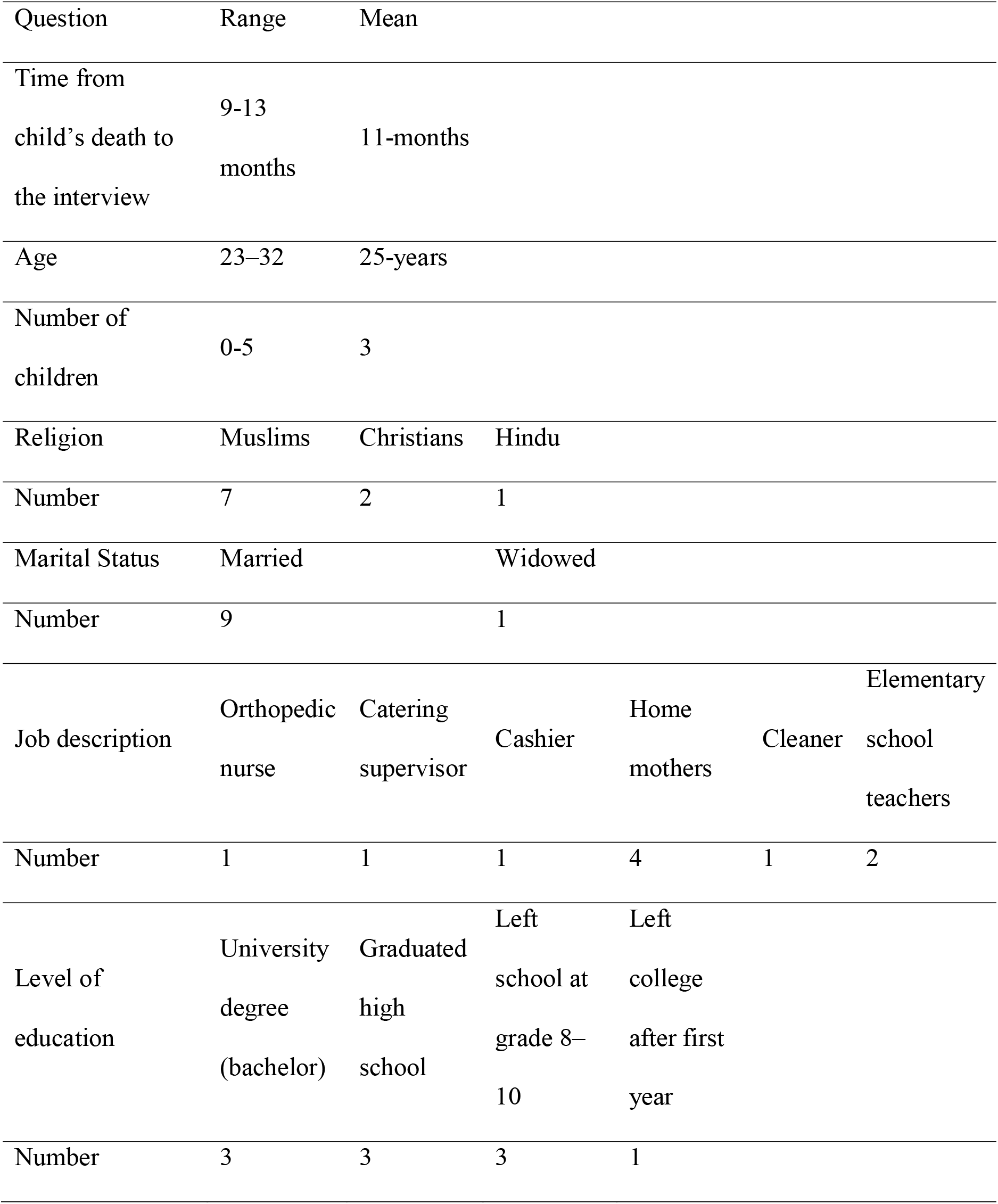
Demographic Questions.

**Table 2:**
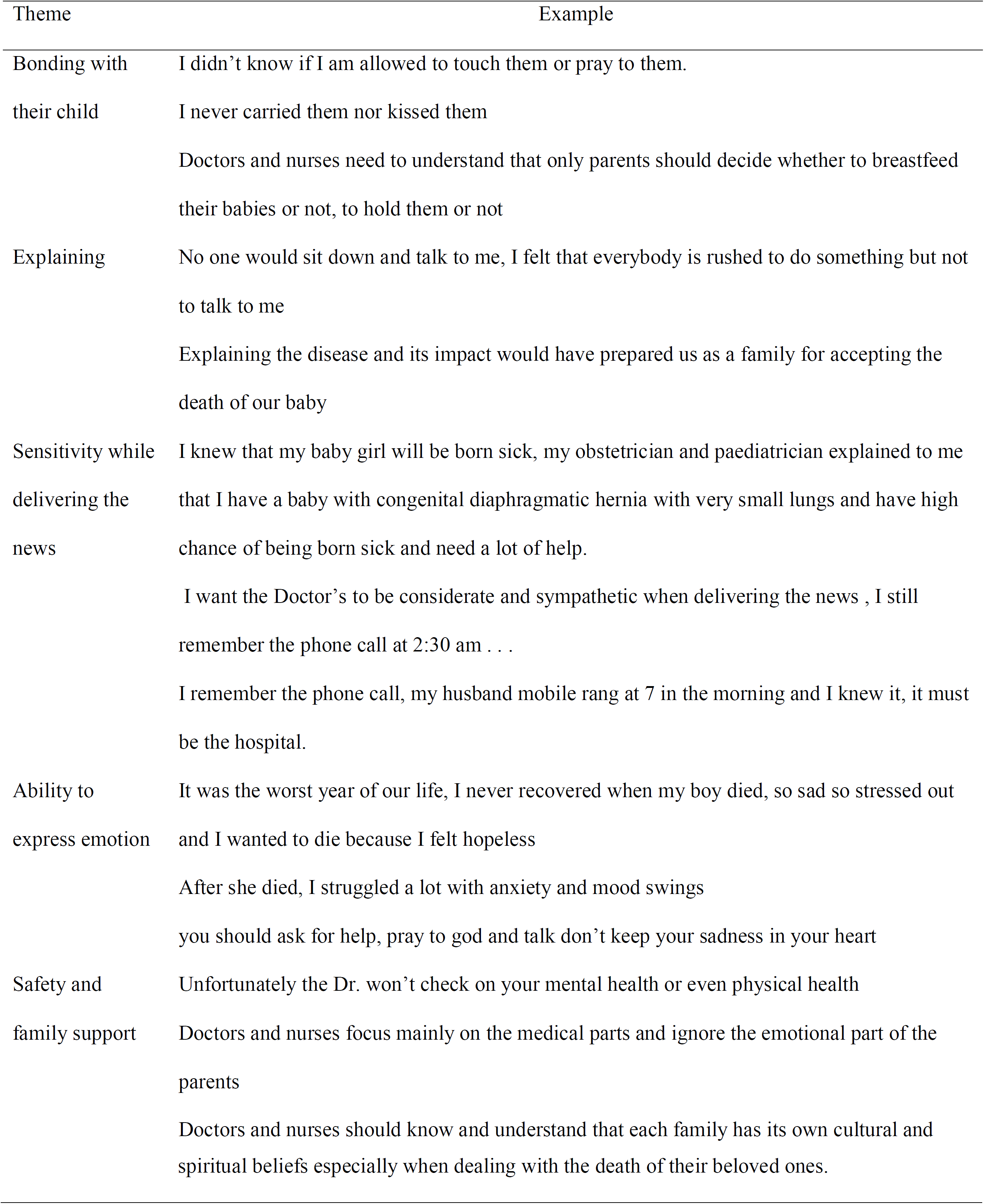
Emerging themes describing mother’s bereavement after neonatal death.

### Theme 1: Bonding with their child

A couple of mother’s regretted not being to spend time with their baby leading to feelings of failure in fulfilling their maternal instincts “I never carried them not kissed them “Another mother also mentioned “I didn’t know if I am allowed to touch them or pray to them.” This has led to feelings of guilt, despair, detachment and worthlessness.

### Theme 2: Explaining

Most of the mothers agreed in the following theme that the HCP where not committed when it came to answer questions clearly in regard to their baby which led to feelings of distress, unappreciated and dishonesty within the doctor / patient relationship “No one would sit down and talk to me, I felt that everybody is rushed to do something but not to talk to me” While one mother mentioned that if the HCP team have shown a more humane approach and honesty regarding the death of her baby it would have eased the process for her and will have a long lasting satisfaction with the treating team. “Explaining the disease and its impact would have prepared us as a family for accepting the death of our baby”

### Theme 3: Sensitivity while delivering the news

One mother has expressed appreciation to the HCP when they were honest, professional, compassionate, and empathetic when delivery grave news in a language that is straight forward with non technical information with total transparency and in a timely matter. “I knew that my baby girl will be born sick, my obstetrician and paediatrician explained to me that I have a baby with congenital diaphragmatic hernia with very small lungs and have high chance of being born sick and need a lot of help.”

While other’s have felt that it is the duty of the HCP to acknowledge the depth of sadness and maternal grief that they may face in order to refine their communication skills showing empathy and compassion “I want the Doctor’s to be considerate and sympathetic when delivering the news, I still remember the phone call at 2:30 am …”

### Theme 4: Ability to express emotion

Most of the mother’s agreed that during this difficult time in order for her to process the pain of her dying baby she should be encouraged and supported thru expression of Numbness, Role confusion, Depression, Detachment, Anxiety, Fear, Anger, Regret, Guilt and Sadness in order for her to find resolution and Grace. “It was the worst year of our life, I never recovered when my boy died, so sad so stressed out and I wanted to die because I felt hopeless”, “After she died, I struggled a lot with anxiety and mood swings”, “you should ask for help, pray to god and talk don’t keep your sadness in your heart”.

### Theme 5: Safety and Family support

A couple of mother’s expressed feelings of irritation and upset towards the HCP for mainly taking care of the baby without checking on their psychological or mental wellbeing. “Doctors and nurses focus mainly on the medical parts and ignore the emotional part of all parents” Which caused them to have feelings of abandonment during the grieving process that led to unexpected emotions from shock or anger to disbelief, guilt and profound sadness. The pain of grief has caused physical pain.

While one mother asked the HCP that in the future they should be mindful and understanding when dealing with a grieving family over a dying baby especially when it comes from a different background culturally and spiritually and they should be able to take care of them and treat them with respect despite the differences “Doctors and nurses should know and understand that each family has its own cultural and spiritual beliefs especially when dealing with the death of their beloved ones.”

## Discussion

### Theme 1: Sensitivity delivering the news

Difficulties may arise with the HCP regarding communication with a bereaving parent which manifest in their own insecurities and anxiety, possibly due to inadequate training in communication techniques and care relations [12]. Communication is not a gift but a skill that can be honed and learnt. The relationship between mother and care team is rooted in partnership and professionalism, the communication should be professional, ongoing, honest, empathetic, consistent and transparent.

In our study mothers have repeatedly stressed out the need for a familiar person to deliver the difficult news using straight forward non technical language with clear information in a timely matter that is also conveyed with compassion and care to her feelings. “I knew that my baby will be born sick with a congenital diaphragmatic hernia, my obstetrician and paediatrician explained to me that” another mother stated that “I want the Doctor’s to be considerate and sympathetic when delivering the news, I still remember the phone call at 2:30 am the phone rang with no introduction nothing calling from hospital … your baby is dead …” The duty of the HCP is to educate the mother to anticipate possible complications and reassure her that she won’t be left alone to face this darkness [13]. Thru the literature it has been shown that grieving mother’s want their grief to be acknowledged and delivering of any information of their baby with total transparency and empathetic information [14, 15]. Although fetal death is common, HCP tend not to listen to their selves while breaking news to the mother which leads her to be emotionally distant. This inability to communicate leads to distrust, conflicts and dissatisfaction with the Quality of care which shows the importance of formal training in breaking bad news since it’s a skill to be learnt [12, 16, 17].

### Theme 2: Bonding with their Child

When faced with the death of a baby, mothers have a difficulty of recognizing the loss. Immediately after birth this might be the only opportunity the mother might have to cuddle, kiss, talk to, bath and dress her baby. The HCP needs to understand the needs and opportunity’s of the grieving mother and may play a unique role in which many actions in the NICU or in the labour and birth area that are should be take to facilitate the bonding attachments of the mother with their baby to produce memories after a sudden or unexpected death that may last for a lifetime. Forming an identity for the baby while it’s alive is an important part of this process and can be accomplished by using the infant’s correct sex and first name, talking to the mother about her baby likes and dislikes and describing their personality. Holding should be facilitated as soon as possible. Mother’s should be encouraged to breastfeed and do kangaroo care when the chance comes. Finally visiting policies should be around the clock, including during bedside rounds so mothers may have the chance to form a strong relationship with HCP team [7, 18, 19].

Thru the analysis it was noted that in multiple occasions mother’s mentioned the need to hold their baby at the time. Having the ability to breastfeed or do kangaroo care. This desire of wanting to fulfil their maternal instincts have been mentioned in the literature having a positive impact on maternal health [20]. The most recent evidence have suggested that ‘mothers feel more natural, good, less frightened and uncomfortable when they see and hold their baby’ [21].

“Doctors and nurses need to understand that only parents should decide whether to breastfeed their babies or not, to hold them or not. Once everyone agreed to go through palliative care path, mothers’ opinions and rights should be respected”

It has been shown thru the analysis negative feelings expressed by mother’s of strong regret and anger at not having the opportunity of being offered to hold her baby to say good bye or spend time with them, which led these mother’s to have feelings of being lost which also has been found in the literature [22].

“There are so many things that I regret or feel sorry about. I didn’t have a chance to carry my baby or feed her. My dream was to carry a baby of my own and breastfeed, and when I had a baby I wasn’t allowed to do so. I didn’t take a picture of her or foot print or piece of her hair. For all mothers who have babies in NICU, don’t wait for the doctors or nurses to offer holding or touching your babies, you do it is your right and you deserve it. Doctors and nurses focus mainly on the medical parts and ignore the emotional part of the parents”.

It has been proven by some well designed descriptive studies that under the right circumstances and guided by compassionate, sensitive and experience staff members mother’s may be able to have the positive experience during this difficult time [23]. During this time of extreme vulnerability of the mother and the difficulty of the situation the HCP have the opportunity to guide the mother following the help of the latest NICE guideline that have undergone many qualitative and quantitative research, has advice that the HCP are in the perfect position to have the mother’s have an interaction with their baby in order to enhance her maternal instincts and help her with the grieving process. The HCP should gradually discuss with the mother what she like to do, support her and encourage her in seeing and holding her dying baby, gather mementos and photographs of the baby [19, 23].

### Theme 3: Safety and Family Support

Mother’s have pointed out the importance of the HCP in taking care of them during this time of vulnerability and how it may give a positive impact to the grieving process. In the following study mothers have reported feelings of anger and frustration when the HCP were insensitive to their feelings and where not checking on them. As stated by one of the mother’s “Unfortunately the Dr. won’t check on your mental health or even physical health … “From the following statement it has been perceived by mothers that they don’t receive the quality of care that they desired. It has been shown in the literature that mother’s expect that the HCP are competent enough, highly trained and skilled in facilitating and providing individualized support to maternal needs [24, 25]. The HCP can make a positive difference in how the death is experienced by the mother by recognizing the impact of death, minimize uncertainty and isolation by thoughtfully working closely with the mother and providing guidance and support by the HCP showing willingness to communicate and care to mother’s [4, 26, 27]. There is a lack of clear clinical guidelines and formal education for the HCP which may complicate the provision of skilled and compassionate care. Which has deemed upon the HCP responsibility to assume a discussion regarding the available resources and support groups that may be provided even though it may change over time requiring anticipatory guidance provided by the HCP and for program planning to ensure community resources that reflect the changing maternal needs [28]. The best Quality of Health Care is shaped by the experiences and engagement of the mothers with the HCP.

### Theme 4: Ability to express emotion

Months of planning and preparation goes into the births and delivery of a new baby. When an unexpected death of a baby happens all these expectations are shattered. It’s difficult to define the normal process of grief which are a bundle of emotions, Grieving response is dynamic pervasive and highly individualized. The other thing is that the process of grief is not linear and doesn’t fit neatly into predetermined categories. As thru the analysis it was shown that many mothers suffer from intense feelings of sadness, despair, helplessness, loneliness, anger, guilt, depression and anxiety. This experience of shock because their loses are overwhelming to the mind and they are normal emotions in the early stages of death. In theory, the strength and severity of separation distress reveals the importance of attachment between mothers and their deceased infants [29].

“When she was born, she was rushed to NICU, she was connected to so many wires and tubes and I knew she will not live. After she died, I struggled a lot with anxiety and mood swings”

It has been proven thru the literature countless of times that these emotional reactions experienced by mothers will have a long term adverse effect on her health if not tackled early on [23]. These emotions during the time may affect the small chance of bonding with the baby having the mother experience intense grief after the death [20].

“It was the worst year of our life, I never recovered when my boy died, so sad so stressed out and I wanted to die because I felt hopeless. Am 44 years old and it is really difficult to get pregnant at this age”

Through the literature, many mothers agreed to experiencing strong emotions that lead to self blame, disappointment in the lost of not being able to fulfill their dreams, fear of repeated pregnancy loss due to the advancement of age. This makes women perceive their loss as being unsuccessful in motherhood [30]. It is well known of the Kubler Ross / Five stages of grief paradigm of Denial, Anger, Bargaining, Depression and acceptance are part of the framework that makes our learning to live with the one lost. The HCP may be able to tackle maternal grieving process using this framework but needs to understand that grief doesn’t have a linear timeline [31]. These stages are responses to loss that many people have but there is not a typical response to loss as there is no typical loss which the HCP should have in mind. Another note in literature is that the staff has to be clear on what the hospital has available by ways of support. Clergy; social workers; pamphlets and books on grieving; and answering medical questions are all important options to offer the grieving mother [24].

### Theme 5: Explaining, Give understanding and meaning

Losing a child has been compared to an amputation of an extremity it is a permanent loss of a part of one self to which one may adjust but which will never return [32]. The literature has shown that families often regard the Health care providers as a second family and thus the lack of contact with staff member’s after a child’s death can be experienced as a “secondary loss”, The HCP is in a position to help the family members cope, both with the immediate loss and then the ongoing effect of the child’s death. Mother’s during the time of loss requires support from the health professionals as well as social and religious support [30]. Mother’s in the literature thrive to have medical attention but won’t ask for it, here the HCP plays the role of providing guidance to these mothers as was shown in the analysis of the need for this interaction

‘Unfortunately, the doctor will not check on your mental health or even physical health. My breast was engorged and full of milk; I didn’t know that I could take tablets to stop the milk from coming!’

“Doctors and nurses should know and understand that each family has its own cultural and spiritual beliefs especially when dealing with the death of their beloved ones”

Given the complexity of supporting bereaved families. It is important to provide education and guidance for Health Care providers, to promote self awareness, therapeutic efficacy, management of professional boundaries and healthy coping mechanisms [33, 34]. The HCP provider should be able to follow up with mothers after the death of her baby, prepare her for what to expect at the time of death including the funeral arrangements and that they should offer support to talk to the HCP whenever possible, psychological support services or support groups with other bereaved parents. It has shown that parents describe a great appreciation towards the HCP when they show basic act of kindness like keeping in touch or attending the funeral [35].

## Conclusions

The death of an infant is a severe loss, and it is important to acknowledge families’ appropriate need to grieve for their babies. Physicians play an important role in the mother’s transition throughout the grief process. Parents do expect the healthcare providers to be competent, be highly trained and skilled in facilitating and supporting parental needs. The key components that should be considered in supporting bereaved parents include honesty, information, choices, and timing. The current study establishes the need for a meaningful and appropriate care for parents and families after the death of their infant.

## Data Availability

Available upon request

## Declarations

### Ethics approval and consent to participate

The informed consent was obtained verbally from all individual participants included in the study.

### Consent for publication

Not applicable

### Availability of data and materials

Not applicable

### Authors’ contributions

AE, AKAA and MK collected and interpreted the patient data. MK performed the data analysis and was a major contributor in writing the manuscript. All authors read and approved the final manuscript.

## Acknowledgements

Not Applicable

## Abbreviations

NICU: Neonatal Intensive Care Unit.

